# Utilization of Cycle Threshold Values of RTPCR SARS-Cov-2 Among Admitted Patients in a Public Hospital

**DOI:** 10.1101/2022.12.05.22283130

**Authors:** Arbeen Acosta Laurito, Alyanna Mae Cuerquis Arengo

## Abstract

Coronavirus 2019 (COVID-19) was caused by the novel severe acute respiratory syndrome coronavirus-2 (SARS-CoV-2) and not just affected the Philippines but also globally. Data on real-time PCR cycle threshold (Ct) values were accessible in the healthcare facility and previous studies did not support its direct clinical significance on patient’s management. The aim of this study was to investigate the RT-PCR Ct values of admitted COVID-19 confirmed patients in the epidemiologic context from April 2021 to November 2021.

A total of 1,245 were tested and out of the 1,038 confirmed COVID-19 admitted patients, 579 (55.78%) were females and 459 were males (44.22%). There were 4 genes detected, namely: N, E, ORF1ab and RdRP genes, and the majority was N gene 925 (88.94%) while the least detected was 203 E gene of SARS-CoV-2. This study described the utilization of five (5) different COVID-19 test kits.

Comparing the Ct values between the male and female groups, this study showed no significant differences. To compare between the different age groups and the three classifications of CT values was shown in Table 6. For the N gene, the current study showed a significant difference of CT value <25 among 0-17 years old vs 46-60 years old (p=0.00054), and among 0-17 years old vs >61 (senior citizens) years old group (p=0.00945). Moreover, a significant difference was observed for CT value >30 among 0-17 years vs 18-45 years old (p=0.00411) and among 0-17 years old vs >61 years old group (p=0.00025). CT values for the ORF gene showed significant differences. CT value <25 showed significant differences among 0-17 years versus 45-60 years old (p=0.00907) and 0-17 years versus >60 years old (p=0.04298). Moreover, CT value >30 showed significant differences among 0-17 years versus 46-60 years old (p=0.02344) and 0-17 years versus >60 years old (p=0.3411).

Comparing the mean CT values of two consecutive months from April to Nov, this study showed a significant difference between April and May (p value= 0.0004), August and September (p value= 0.0212), and September and October (p value= 0.002). There was no significant difference with the following months: May and June, June and July, July and August, and October and November (Table 9).

## INTRODUCTION

Late in the year 2019, a new corona virus was identified. Subsequently, early in the year 2020, an outbreak was declared in Wuhan, China which later spreading to the neighboring countries into a pandemic. (1) As the pandemic continued, different variants arose (alpha, beta, delta, epsilon, eta, lota, kappa, 1.617.3, mu, zeta, omicron). (2) Among infected individuals, there was a wide range of clinical manifestations of the SARs COV infection, some presenting as asymptomatic, or some with fever, cough, colds to shortness of breath to acute respiratory syndrome, generally classified as mild cases to severe and from each spectrum there were different outcomes. (3)

According to Center for Disease Control and Prevention (2022), different variants have different transmissibility and have different outcomes. There are variants that were more transmissible but present as mild and less transmissible but could have detrimental outcomes. (4) As of the present there has been a total number of 3.69 million COVID cases and 60,455 deaths in the Philippines. (5) Studies are being done to establish the risk factors affecting the infectivity and outcomes such as demographics, epidemiology and diagnostics

One of the strategies to control the spread of the virus and prevent progression of illness was testing. Generally, there were 3 kinds of test: Serologic, nucleic acid amplification test and RT PCR test, which was considered to be the gold standard being fast, specific and sensitive to the virus. (6) Part of the quantitative measure of the test is the cycle threshold value which was the number of cycles needed for an amplicon to be detectable above background. Lower CT values indicated a higher viral load and a higher CT-values indicated lesser viral load thus lesser infectivity. Predicting the infectivity, in some institutions, this has been used as the basis for clinical decisions to be made.

This is a cross-sectional study aims to assess the admitted patients diagnosed with COVID 19 based on demographics, evaluated the kits used, and the specific gene results from the RT PCR.

### Objectives of the study

CT values have limitations for use in the clinical settings. Low CT values means high transmissibility of viral load, and high CT values would mean a low transmissibility of the COVID-19 virus. These CT values were used as the gold standard in diagnosing the admitted patients in the hospital through a platform-base PCR. The absence of published study in the local setting utilizing these available data made the researchers exploit and analyze existing CT values among the admitted patients at a city-wide level.

### Specific Objectives

1. To describe the distribution and differences of CT values among COVID-19 admitted patients in terms of CT value results, sex, and different age groups.
2. To determine the presence and absence of COVID-19 detectable genes and its differences with coexistence of other genes.
3. To describe the distribution of CT value results based on the utilized COVID-19 test kits among COVID-19 admitted patients.
4. To determine the differences of the monthly CT values among COVID-19 admitted patients.

### Scope of the study

The current study included all COVID-19 admitted patients who were city residents, regardless of age, initial signs and symptoms, and their clinical outcomes. Results of RT-PCR were based on an anonymized master list from the healthcare facility. There four (4) genes utilized in the study based on the detected genes among the used test kits. Three sub-groups of the viral load (namely: <25, 25-30, >30) were used as the basis for distribution among other parameters used in the study.

### Limitations of the study

This study is a document review on CT values results from hospital records. There was no patient interaction and no control on the timing of swabbing and area to be swabbed. There was also no control on the type and supply of test kits used. Clinical status of the patients (asymptomatic or symptomatic) was not determined during the conduct of the study. All specimens were sent to a single accredited referral molecular laboratory. Results of the RT-PCR were received by the facility as batches containing the positive (confirmed COVID-19 infection), negative and equivocal results (for repeat swabbing).

## METHODOLOGY

The study was a purposive cross-sectional study design utilizing anonymized hospital records of platform-based RT-PCR CT values among admitted COVID-19 confirmed patients from April to November 2021 in a level-I government hospital.

The CT values were collated from anonymized hospital records during the study period after the approval of the hospital ethics committee and the Chief of Hospital. The study commenced last April 2021 since the hospital started to access the data from the external molecular laboratory and hospital database was functional at that time. The December 2021 data were excluded since detected cases were much lower compared to April 2021 and some were incomplete.

The clinical criteria to qualify for swabbing, timing when to do swabbing from the asymptomatic to symptomatic status, the choice of doing an oropharyngeal or nasal swabs per patient, and the staff who performed swabbing were beyond the scope of the study. All viral samples were sent out to a referral molecular laboratory. Based on the record, the study population was selected only among residents of the city, presence of CT values, and admitted to the hospital. Since it was a record review, no actual and direct communication with the patients was held and there was no follow-up with their clinical status.

City residents who were confirmed COVID-19 infection but admitted at the step-down facilities or those referred to apex tertiary hospital were excluded in the study.

Sex of the study population were limited to male and female, while age groups were as follows: 0-17 years old (pediatric group), 18-45 years old, 45-60 years old, and >60 years old (senior citizens population). This age group was correlated to the series of vaccination roll out in different age groups as strategized by the Philippine Department of Health (DOH).

Results of the CT values were used as the “gold standard” in determining a confirmed COVID-19 infected admitted patient. The researchers did not focus on specific test kits and had no control on the use of a specific test kit since the hospital and region had challenging experience with the supply and demand of these test kits. During the course of the study, the external molecular laboratory utilized 5 different test kits, namely; 1) Biorad (Bio-Rad SARS-CoV-2 ddPCR Kit), 2) Fortitude test kit (Fortitude Kit), 3) Genefinder (GeneFinder™ COVID-19 Plus RealAmp Kit), 4) Sansure (Novel Coronavirus (2019-nCoV) Nucleic Acid Diagnostic Kit [PCR-Fluorescence Probing]), and 5) Solgent (SolGent DiaPlexQ™ PCR Test Kit). Consolidated CT values were grouped into three according to the level of detection and sensitivity of the test kits, as follows; low CT value <25 (high viral load), moderate CT value of 25-30 (intermediate viral load), and high CT value>30 (low viral load).

COVID-19 test kits were utilized invariably during the study period. The results of RT-PCR among the study population also invariably presented the detected specific gene or genes at different viral loads. Moreover, this study described the target genes detected which were dependent on the sensitivity and specificity of the test kits used, as follows: N gene, ORF gene, (gene symbol ORF1ab: Gene description: RF1a polyprotein; polyprotein) (NLM, 2022), RDRP gene (RNA-dependent RNA polymerase gene) (Cho et al., 2020), and E gene of SARS-CoV-2.

There were two RT-PCR machines used, namely Applied Biosystems™ 7500 Real-Time PCR Systems by Thermo Fisher Scientific Inc. and Bio-rad CFX Opus 96 Real-Time PCR Instrument (with 96-well, network-connected real-time PCR detection system) by Bio-Rad Laboratories, Inc. which were both utilized simultaneously or individually during the study period.

Anonymized data were retrieved from the hospital records. Collated CT values were regrouped accordingly to address the specific objectives of the study. These were presented in terms of frequency, mean, range, percentage and according to the viral load association as follows; low CT value <25 (high viral load), moderate CT value of 25-30 (intermediate viral load), and high CT value>30 (low viral load). Study population sex were grouped into male and female.

The CT values distribution were also described in terms of the gene detected and test kits being utilized in terms of frequency, mean and percentage. In some detected genes, the majority of the study population would have no CT values and were considered as ‘not detected’. Thus, these were not subjected to any qualitative analysis.

The data collected were analyzed using two-tailed unpaired t-test at 95% confidence interval (alpha 0.05). These were presented in terms of standard deviations (SD), confidence interval, t value, standard error of difference and the p value. Data with frequency less than 5 were not subjected to any statistical analyses.

## RESULTS

Table 1 presented the distribution of COVID-19 genes using the plate-based RT-PCR among 1038 COVID-19 admitted patients as study population, namely: N gene, ORF gene, RDRP gene and E gene, with their respective frequency, mean CT value and range of CT values. In addition, CT value was subclassified into 3, as: <25-high viral load, 25-30 as intermediate viral load, and >30 low viral load.

**Table 1.** The frequency, and the mean and range of CT values among COVID-19 admitted patients. (n=1040)

| Gene | Parameters | CT value |  |  | Total | % |
| --- | --- | --- | --- | --- | --- | --- |
|  |  | <25 | 25-30 | >30 |  |  |
| N gene | Frequency | 343 | 208 | 347 | 925 | 88.94 |
|  | Mean CT value | 20.56 | 27.70 | 35.65 |  |  |
|  | Range CT value | 12.36-25 | 25.03-30 | 31-43.28 |  |  |
| ORF | Frequency | 265 | 145 | 336 | 746 | 71.73 |
|  | Mean CT value | 21.046 | 27.42 | 35.640 |  |  |
|  | Range CT value | 12.0-25 | 25.01-30 | 30.04-41.9 |  |  |
| RDRP | Frequency | 55 | 69 | 90 | 214 | 20.58 |
|  | Mean CT value | 21.50 | 27.628 | 35.747 |  |  |
|  | Range CT value | 12.15-25 | 25.03-30 | 30.16-44.747 |  |  |
| E gene | Frequency | 70 | 59 | 74 | 203 | 19.52 |
|  | Mean CT value | 21.04 | 27.54 | 34.62 |  |  |
|  | Range CT value | 12.51-25 | 25.267-30 | 30.02-42.603 |  |  |

The majority of these admitted patients had N gene detected with 925 or 89% of the study population. Furthermore, most of these patients had CT value >30 with 347 patients (mean CT value=35.65), followed by <25 (mean CT value=20.56), then 25-30 CT value (mean CT value=27.70). The ORF gene was detected in about 746 patients (71.73%), 214 patients RDRP gene (20.58%), and 203 E gene detected admitted patients (19.52%). This study consistently documented that the majority of admitted patients detected COVID-19 specific genes with CT values (see Table 1).

The mean CT values and range of CT values were presented in each gene relative to its CT value groups. The frequency and the mean CT values among male COVID-19 admitted patients (n=459) were presented in Table 2 with different genes and CT value groups. The N gene was detected 88% (404) among 459 male COVID-19 admitted patients. It was followed by ORF gene (72.77%), RDRP gene (20%), and the least was E gene (18.74%).

**Table 2.** The frequency and the mean CT values among **male** COVID-19 admitted patients (n=459).

| Gene | Parameters | CT value |  |  | Total | % |
| --- | --- | --- | --- | --- | --- | --- |
|  |  | <25 | 25-30 | >30 |  |  |
| N gene | Frequency | 152 | 91 | 161 | 404 | 88 |
|  | Mean CT value | 20.87 | 27.31 | 35.45 |  |  |
| ORF gene | Frequency | 120 | 70 | 144 | 334 | 72.77 |
|  | Mean CT value | 21.05 | 27.42 | 35.59 |  |  |
| RDRP gene | Frequency | 20 | 34 | 38 | 92 | 20 |
|  | Mean CT value | 21.731 | 27.57 | 35.35 |  |  |
| E gene | Frequency | 28 | 29 | 29 | 86 | 18.74 |
|  | Mean CT value | 21.88 | 27.21 | 34.42 |  |  |
| **Total** | **Frequency** | **148** | **104** | **207** | **459** | **100** |
|  | **Mean CT value** | **20.99** | **27.32** | **35.67** |  |  |

Based on the CT values, the majority of the N gene among male COVID-19 admitted patients had 161 (>30) for N gene, then 152 for <25, and 91 admitted patients for the range of 25-30. The observation that the majority had >30 CT value were consistent for other genes, namely: ORF gene 144/334; RDRP gene 38/92, and 29/86 for E gene.

The N gene was detected 90% (520) among 581 female COVID-19 admitted patients. It was followed by ORF gene (71%), RDRP gene (21%), and lastly, E gene at 20% (Table 3).

**Table 3.** The frequency and the mean CT values among female COVID-19 admitted patients (n=581).

| Gene | Parameters | CT value |  |  | N | % |
| --- | --- | --- | --- | --- | --- | --- |
|  |  | <25 | 25-30 | >30 |  |  |
| N gene | Frequency | 190 | 98 | 232 | 520 | 89.5 |
|  | Mean CT value | 20.30 | 27.49 | 35.37 |  |  |
| ORF | Frequency | 144 | 75 | 192 | 411 | 70.74 |
|  | Mean CT value | 21.02 | 27.41 | 35.68 |  |  |
| RDRP | Frequency | 35 | 35 | 52 | 122 | 21 |
|  | Mean CT value | 21.61 | 27.68 | 35.16 |  |  |
| E gene | Frequency | 42 | 30 | 45 | 117 | 20 |
|  | Mean CT value | 20.48 | 27.85 | 34.75 |  |  |
| Total | Frequency | 191 | 100 | 290 | 581 | 100 |
|  | Mean CT value | 20.72 | 27.46 | 35.64 |  |  |

Based on the CT values, the majority of the N gene among female COVID-19 admitted patients had >30 (232), then <25 (190), and 25-30 with 98 patients. These observations that the majority had >30 CT values were similar to other genes, namely: ORF gene 192/411; RDRP gene 52/122, and 45/117 for E gene.

The pooled distribution of detected and non-detected N gene, RDRP, ORF, and E genes between male and female admitted COVID-19 patients are shown in Table 4 relative to the CT values groupings, as follows: < 25, 25-30, and >30.

**Table 4.** Distribution of both male and female sexes relative to different genes and corresponding CT values.

| Gene | Parameter | Male |  | Female |  |
| --- | --- | --- | --- | --- | --- |
|  |  | n | Total | n | Total |
| N gene | < 25 | 151 | 403 | 191 | 520 |
|  | 25-30 | 91 |  | 98 |  |
|  | >30 | 161 |  | 231 |  |
|  | Not detected | 55 | 55 | 60 | 60 |
| RDRP gene | < 25 | 20 | 92 | 35 | 121 |
|  | 25-30 | 34 |  | 35 |  |
|  | >30 | 38 |  | 51 |  |
|  | Not detected | 366 | 366 | 459 | 459 |
| ORF | < 25 | 119 | 333 | 146 | 411 |
|  | 25-30 | 70 |  | 73 |  |
|  | >30 | 144 |  | 192 |  |
|  | Not detected | 125 | 125 | 169 | 169 |
| E gene | < 25 | 28 | 86 | 42 | 116 |
|  | 25-30 | 29 |  | 30 |  |
|  | >30 | 29 |  | 44 |  |
|  | Not detected | 372 | 372 | 464 | 836 |

For the N gene, the majority of the male (161) and female (231) admitted patients had CT value >30. These distributions were followed by populations with CT values <25 and 25-30 for both male and female populations. The same pattern was observed for RDRP gene (male 38 (>30), female 51 (>30)], ORF gene (male 144 (>30), female 192 (>30)], and E gene (male 29 (>30), female 44 (>30)].

The minority of the population for each gene were observed with CT value 25-30, as follows: N gene both male (91) and female (98) sexes while ORF gene with 70 for male and 73 for females. In contrast, a minority of the population were observed with variable CT value distribution for RDRP gene (male=20 CT value <25, female=35 both CT values <25 and 25-30), and E gene (male=28 CT value <25, female=30 CT value 25-30).

The distribution of detected and non-detected CT values for N gene, RDRP, ORF, and E genes between male and female admitted COVID-19 patients were shown in Table 5.

**Table 5.**
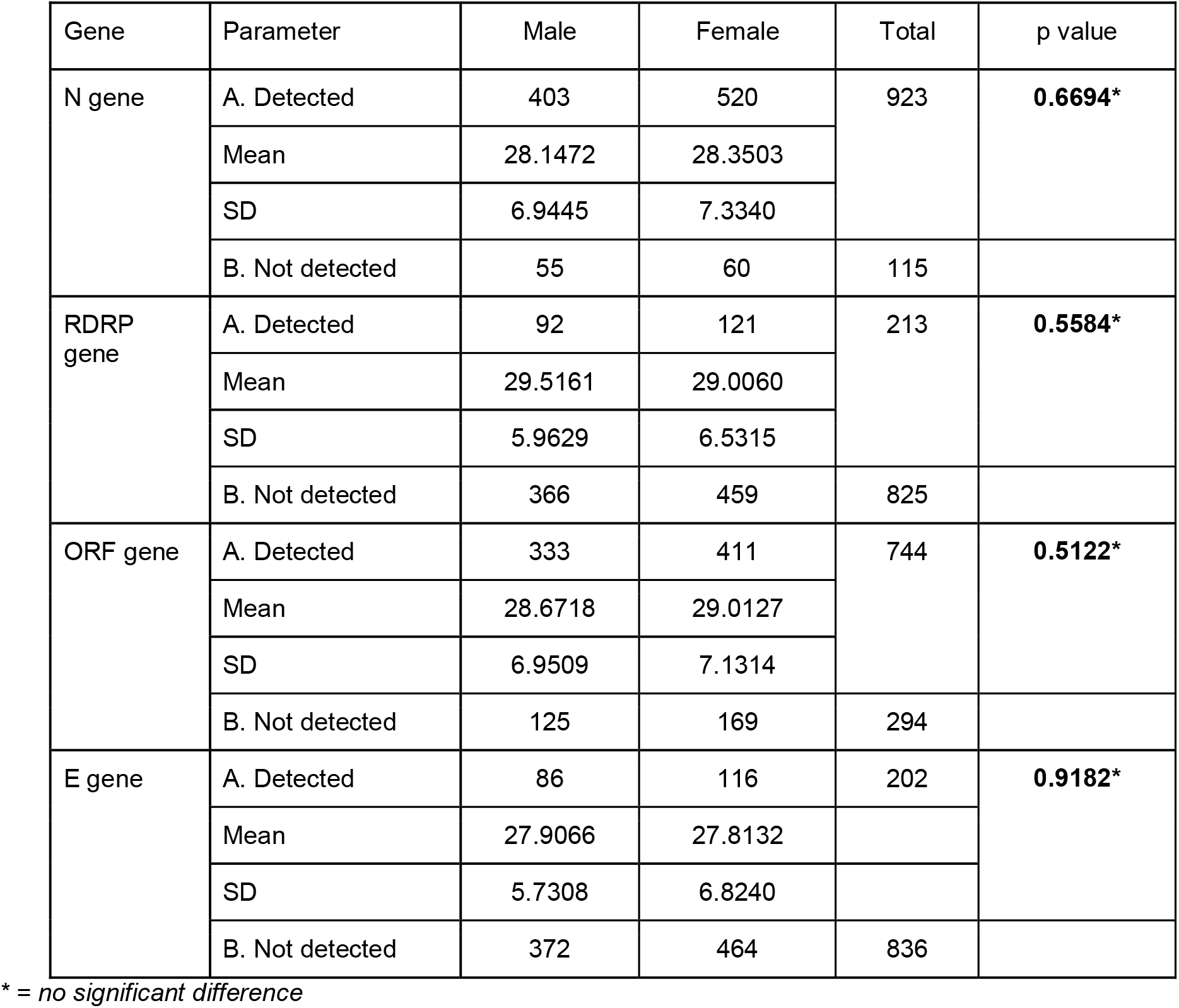
The distribution of detected and non-detected CT values for different genes between male and female admitted COVID-19 patients.

**Table 6.** The distribution of different age group relative to the different genes and CT values among COVID-19 admitted patients.

| Gene | Parameter | Age group (years) |  |  |  | P value |  |
| --- | --- | --- | --- | --- | --- | --- | --- |
|  |  | T1 | T2 | T3 | T4 |  |  |
|  |  | 0-17 | 18-45 | 46-60 | >61 |  |  |
| Frequency |  | 92 | 449 | 239 | 258 |  |  |
| N gene | < 25<br>(Mean CT value) | 17<br>19.0128 | 161<br>20.0034 | 78<br>21.5045 | 86<br>21.0014 | 0.0002289* | T1:T3 p=0.00054<br>T1:T4 p=0.00945 |
|  | 25-30<br>(Mean CT value) | 7**<br>** | 79<br>27.4400 | 48<br>27.2147 | 55<br>27.3815 | 0.725402 |  |
|  | >30<br>(Mean CT value) | 49<br>36.9671 | 159<br>35.4936 | 88<br>34.6352 | 96<br>35.1735 | 0.000054* | T1:T2 p=0.00411<br>T1:T3 p=0<br>T1:T4 p=0.00025 |
|  | Not detected | 19 | 50 | 27 | 21 |  |  |
| RDRP gene | < 25<br>Mean CT value | 5** | 27<br>21.6183 | 8<br>21.459 | 15<br>21.0898 | 0.18331 |  |
|  | 25-30<br>Mean CT value | 3** | 22<br>27.469 | 25<br>27.7371 | 19<br>27.5502 | 0.811457 |  |
|  | >30<br>(Mean CT value) | 5** | 36<br>35.6343 | 17<br>34.3557 | 31<br>34.9277 | 0.4621 |  |
|  | Not detected | 79 | 364 | 189 | 193 |  |  |
| ORIF gene | < 25<br>(Mean CT value) | 13<br>19.6931 | 129<br>20.6854 | 57<br>21.7771 | 67<br>21.4283 | 0.012941* | T1:T3 p=0.00907<br>T1:T4 p=0.04298 |
|  | 25-30<br>(Mean CT value) | 5** | 68<br>27.6001 | 35<br>27.0474 | 34<br>27.4247 | 0.211627 |  |
|  | >30<br>(Mean CT value) | 52<br>36.541 | 135<br>35.7096 | 78<br>35.2352 | 71<br>35.2958 | 0.043566* | T1:T3 p=0.02344<br>T1:T4 p=0.3411 |
|  | Not detected | 22 | 117 | 69 | 86 |  |  |
| E gene | < 25<br>(Mean CT value) | 4** | 33<br>20.5273 | 15<br>22.4078 | 18<br>20.7939 | p=0.168554 |  |
|  | 25-30<br>(Mean CT value) | 1** | 27.4977 | 19<br>27.5725 | 21<br>27.5495 | p=0.0984817 |  |
|  | >30<br>(Mean CT value) | 3** | 33<br>35.3049 | 16<br>33.7925 | 21<br>34.2041 | p=0.242286 |  |
|  | Not detected | 84 | 365 | 189 | 198 |  |  |

Note: \* = statistically significant \*\* = not subjected to statistical tool due to small in frequency

Among this study population, the following genes were mostly detected for both male and female population: N gene (923) and ORF gene (744). RDRP and E genes were detected least among male and female patients, with 213 and 116 patients, respectively.

Comparing the occurrences of detected genes between the male and female population, the current study showed no significant differences between their CT values for N gene (p-value=0.6694), RDRP gene (p-value=0.5584), ORF gene (p-value=0.5122), and E gene (p-value= 0.9182).

Moreover, Table 5 presented the mean CT values and standard deviations among the detected genes for both sexes.

The study population were re-group into four groups accordingly, as follows: 0-17 years old (n=92), 18-45 years old (n=449), 46-60 years old (n=239) and >61 years old (n=258), which are currently applied for a population-based category in the community. (See Table 6)

To compare between the different age groups and the three classifications of CT values was shown in Table 6. For the N gene, the current study showed a significant difference of CT value <25 among 0-17 years old vs 46-60 years old (p=0.00054), and among 0-17 years old vs >61 (senior citizens) years old group (p=0.00945). Moreover, a significant difference was observed for CT value >30 among 0-17 years vs 18-45 years old (p=0.00411) and among 0-17 years old vs >61 years old group (p=0.00025).

Table 6 illustrated the CT values for the ORF gene the significant differences. CT value <25 showed significant differences among 0-17 years versus 45-60 years old (p=0.00907) and 0-17 years versus >60 years old (p=0.04298). Moreover, CT value >30 showed significant differences among 0-17 years versus 46-60 years old (p=0.02344) and 0-17 years versus >60 years old (p=0.3411). There was no significant difference comparing four age groups relative to the CT value of 25-30.

On the other hand, this study showed no significant differences among the CT values of RDRP gene when compared among the four age groups, as follows: <25 (p=0.18331), 25-30 (p=0.811457), and >30 (p=0.4621).

CT values for the ORF gene showed significant differences. CT value <25 showed significant differences among 0-17 years versus 45-60 years old (p=0.00907) and 0-17 years versus >60 years old (p=0.04298). Moreover, CT value >30 showed significant differences among 0-17 years versus 46-60 years old (p=0.02344) and 0-17 years versus >60 years old (p=0.3411). There was no significant difference comparing four age groups relative to the CT value of 25-30.

All E gene CT values showed no significant differences compared among all four age groups of COVID-19 admitted patients (see Table 6). The E gene of the majority of these patients was not detected by the available test kits during the study period. CT values for 0-17 years old were not subjected to the statistical tool due to its small numbers.

The presence of CT values in each gene detected were the only values subjected to statistical analysis. Therefore, the frequency of absent CT values was not calculated. The coexistence of each gene was compared to test the significance difference on its CT values between N genes, ORF, RDRP and E genes (Table7).

**Table 7.**
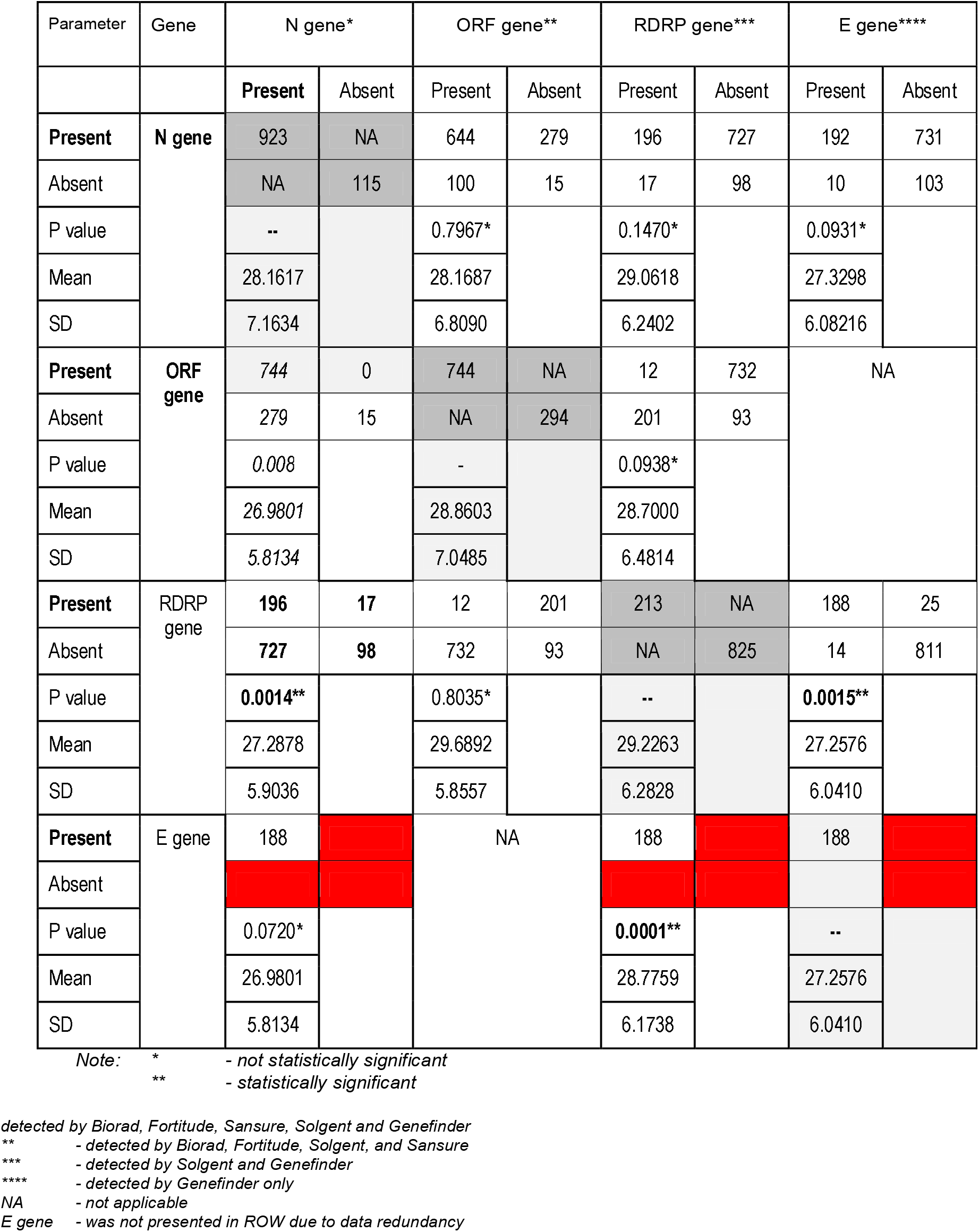
Distribution and comparison of COVID-19 genes coexisting with other genes.

**Table 8.** Coexistence of detected genes using the plate-based RT=PCR.

| Gene | Status | N gene* |  | ORIF gene** |  | RDRP gene*** |  | E gene**** |  |
| --- | --- | --- | --- | --- | --- | --- | --- | --- | --- |
|  |  | Present | Absent | Present | Absent | Present | Absent | Present | Absent |
| N gene | Present | 923 | NA | 644 | 279 | **196** | **727** | 192 | 731 |
|  | Absent | NA | 115 | 100 | 15 | **17** | **98** | 10 | 103 |
| ORIF | Present | 744 | 0 | 744 | NA | 12 | 732 | NA | 744 |
|  | Absent | 279 | 15 | NA | 294 | 201 | 93 | 202 | 87 |
| RDRP gene | Present | **196** | **17** | 12 | 201 | 213 | NA | 188 | 25 |
|  | Absent | **727** | **98** | 732 | 93 | NA | 825 | 14 | 811 |

Note:
\* - Biorad, Fortitude, Sansure, Solgent and Genefinder
\*\* - Biorad, Fortitude, Solgent, and Sansure
\*\*\* - Solgent and Genefinder
\*\*\*\* - Genefinder only
NA - not applicable
E gene - was not presented in ROW due to data redundancy

The N gene (mean CT value= 28.1617) was compared the CT values of the ORF, RDRP and E genes which showed no significant difference based on their p values 0.7967 (mean CT value= 28.1687), 0.1470 (mean CT value= 29.0618), and 0.0931 (mean CT value= 27.3298), respectively.

The ORF gene was detected among 744 patients (mean CT value= 28.8603) was compared the CT values of the same frequency of 744 N and 12 RDRP genes which also showed no significant difference based on their p values *0.008* (mean CT value= 26.9801) and 0.0938* (mean CT value= 28.7000), respectively.

Coexistence of the RDRP gene-detected patients with N genes, ORF, and E gene showed variable results when their CT values were compared. The 213 RDRP gene-detected samples were significantly different with the 196 N gene-positive (0.0014) patients (p value=0.0014) and also with 188 E gene-detected (p value 0.0015) patients. However, CT values when compared to ORF, this showed that there was no significant difference between the two steps of study population as shown in Table 7.

Lastly, CT values of 188 E genes were shown to be significantly different relative to the same frequency of RDRP genes (188, p value=0.0001) while there was no significant difference when compared to 188 N gene CT values (p value= 0.0720).

The majority of the admitted patients had N gene detected (923) out of 1040 study population. Also, other genes detected were ORIF gene, RPRD gene, and E gene, in decreasing order. The N gene was best detected by all of the 5 test kits, namely: Biorad, Fortitude, Sansure, Solgent and Genefinder, in varying viral loads.

Furthermore, Table NN, showed that N gene was detected together with other genes, as follows: RDRP gene in 196 COVID-19 admitted patients, 644 patients together with ORIF gene, and the least with 192 COVID-19 admitted patients together with E gene.

RDRP gene was detected less frequently together with N gene (196 vs 727) and ORIF gene (12 vs 732). This study showed that the RDRP gene was detected by the Solgent test kit and Genefinder. However, R gene was detected in most of the COVID-19 admitted patients together with E gene (188) with Genefinder as the sole test kit for the latter.

The presence or absence of ORIF gene was also observed in this study with the majority of the ORIF gene being detected with N gene (744 patients) while 279 patients had N gene but no ORIF gene. There were only 12 patients with coexisting N and ORIF genes while 732 patients had ORIF gene without RDRP gene. Occurrence of ORIF and E gene together was not detected in this study since ORIF gene was specifically detected by 4 test kits (Biorad, Fortitude, Solgent, and Sansure) other than Genefinder which was more specific for E gene. Moreover, there were 202 patients E gene detected using Genefinder without the coexistence of ORIF gene.

As presented in Table 9, there were five test kits used within the study period. Biorad (Bio-Rad SARS-CoV-2 ddPCR Kit) were used in 14 (1.35%) COVID-19 admitted patients: 2 with CT values <25, 2 for 25-30, and 10 had >30 CT values. Fortitude test kit (Fortitude Kit) used in 30 (2.88%) admitted patients with the majority detected 12 with CT values <25, then 10 patients with >30 CT value, and last was 25-30 with 8 patients.

**Table 9.** Frequency and mean of test kits used among COVID-19 admitted patients relative to the cycle threshold cycles (CT) values. (n=1040)

| Test kits | Parameters | CT value |  |  | Total | % |
| --- | --- | --- | --- | --- | --- | --- |
|  |  | <25 | 25-30 | >30 |  |  |
| 1.Biorad CFX96 | Frequency | 2 | 2 | 10 | 14 | 1.35 |
|  | Mean | 22.05 | 28.39 | 36.93 |  |  |
| 2.Fortitude | Frequency | 12 | 8 | 10 | 30 | 2.88 |
|  | Mean | 18.82 | 26.95 | 34.69 |  |  |
| 3.Genefinder | Frequency | 65 | 68 | 130 | 263 | 25.29 |
|  | Mean | 21.33 | 27.53 | 35.42 |  |  |
| 4.Sansure | Frequency | 72 | 31 | 47 | 150 | 14.42 |
|  | Mean | 20.40 | 27.88 | 34.83 |  |  |
| 5.1 Solgent | Frequency | 182 | 92 | 289 | 563 | 54.13 |
|  | Mean | 20.94 | 27.14 | 35.86 |  |  |
| 5.2 Solgent DuaplexQ | Frequency | 4 | 5 | 11 | 20 | 1.92 |
|  | Mean | 21.38 | 27.90 | 35.91 |  |  |
| Total (Frequency) |  | 337 | 206 | 497 | 1,040 |  |

Genefinder (GeneFinder™ COVID-19 Plus RealAmp Kit) was used in 263 patients (25%) and detected 130 CT value >30, 65 patients with CT value >25, and 68 patients with CT value 25-30. On the other hand, Sansure (Novel Coronavirus (2019-nCoV) Nucleic Acid Diagnostic Kit [PCR-Fluorescence Probing]) detected 150 patients (14%). The majority of these patients (72) had CT value <25, followed by 47 patients who had CT value >30 and 31 with CT value 25-30.

Combining all above test kits, still Solgent surpassed the other four kits. More than (54%) of the admitted patients were tested with Solgent (SolGent DiaPlexQ™ PCR Test Kit) which showed that 289 had CT value >30, followed by 182 patients with CT value <25, and 92 patients with CT value 25-30.

Table 10 showed the comparison of monthly CT values among the COVID-19 admitted patients. The number of admitted patients was lowest at the start (April, 2021) and at the end (November, 2021) within the study period. Furthermore, the peak was observed during the month of August with 373 patients during the COVID-19 surge of the Omicron variant. The month of August also showed the lowest monthly mean CT value of 28.1048 among the COVID-19 admitted patients.

**Table 10.**
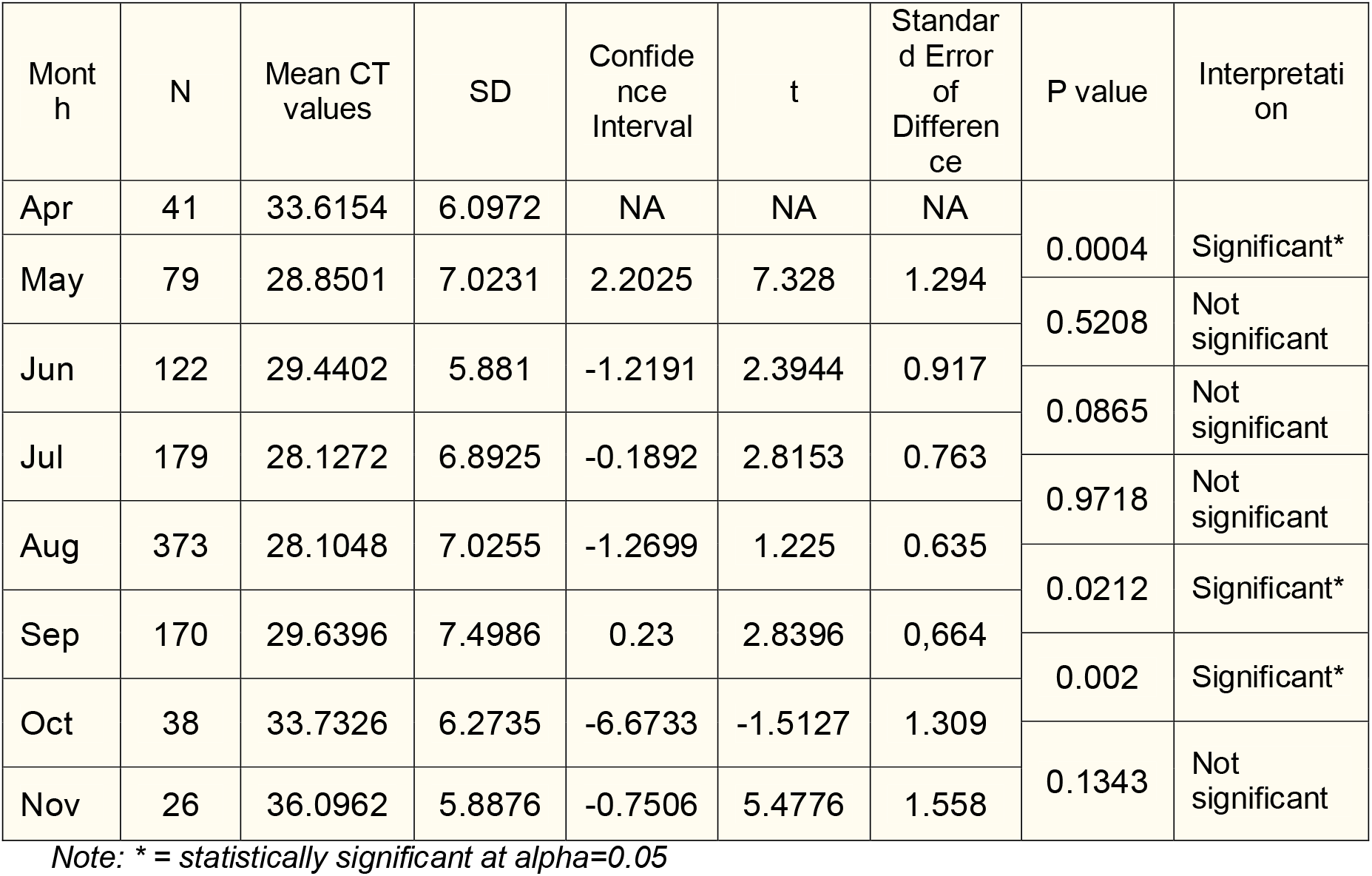
Monthly comparison of the mean CT values of COVID-19 admitted patients.

Comparing the mean CT values of two consecutive months from April to Nov, this study showed a significant difference between April and May (p value= 0.0004), August and September (p value= 0.0212), and September and October (p value= 0.002). There was no significant difference with the following months: May and June, June and July, July and August, and October and November (Table 10).

## DISCUSSION

The study population were group into four sub-groups representing the pediatric age group (0-17 years old), young adult (18-45 years old), older adult (46-60 years old) and the senior citizens (>61 years old).

The relationship between Ct values and age were observed to be the inversely proportional with increasing age (Ibrahin, 2020).

However, this current study showed that Ct values when compared with age per gene had significant differences in the Ct value <25 and >30 for N gene (Ct value <25, p value= 0.0002289 and >30, p value 0.000054) and ORF gene (Ct value <25, p value= 0.012941 and >30, p value 0.043566) (Table 6). Among the 324 samples where virus detection was categorized according to age, Singanayagam et al., (2020) documented no significant difference in Ct values (p= 0.12) and in culture positivity (p= 0.63) from upper respiratory tract samples across the different age groups (0-20, 21-40, 41-40, 61-40, and 81-100 years old).

Kim et al., (2021) investigated the association of viral load dynamics with patient’s age and severity of COVID-19 using a set of respiratory specimens longitudinally collected from 64 patients with different clinical severity and age during acute phase. Higher viral burden, primarily in the younger cohort (≤59 years old) in contrast to the elderly patients (≥60 years old) with critical disease. The study used 10-year age intervals age and smaller study population compared to the current study. Meanwhile, a systematic review of 1337 individuals tested positive for SARS-CoV-2 from RT-PCR tests were associated with risk of mortality in SARS-CoV-2 infection (Waudby-West, 2021). Hazards of Ct values <20 compared to >30 was 2.20 (95% CI 1.28–3.76) in a model adjusted for age, sex, comorbidities and hospitalization.

Chauhan et al., (2021) accepted the possibility that children and older adults (>60 years) have different mechanisms of action against SARS-CoV-2 infection, which needs further study. However, the emergence of escape mutants, infection rate of different age groups, and target genes or proteins for each commercialized kit are different, are the variable considered to be addressed by analyzing large study population.

Levine-Tiefenbrun et al., (2022) analyzed 843,917 test results of 521,696 patients using a single test kit (Allplex 2019-nCoV, Seegene) on quantitative RT-PCR (RT-qPCR). The overall detection rate in tests performed within 14 days after diagnosis was 83.1%, and highest at days 0 to 5 after diagnosis (89.3%). In addition, detection rate was strongly associated with age and sex.

Though within the same range of 25-30 CT value, E gene detection was significantly different than RdRP gene and had higher mean Ct value (p value 0.0001). Test kits used in this study specifically detected 188 patients with both E gene and RdRP and none for 3 genes including N gene. Colton et al., (2020) reported that Sheffield Teaching Hospitals NHS Foundation Trust (UK) experienced on serially sampled patients with confirmed SARS-CoV-2 infection suggested that E gene detection persists beyond RdRp detection, and may offer enhanced diagnostic sensitivity.

Utilizing Ct values for clinical diagnosis still requires more evidences. Population level Ct values was conducted by Walker et al (2021). Of 3,312,159 nose and throat swabs, 27,902 (0.83%) were RT-PCR-positive, 10,317 (37%), 11,012 (40%), and 6550 (23%) for 3, 2, or 1 of the N, S, and ORF1ab genes, respectively. In conclusion, marked variation in community SARS-CoV-2 Ct values suggests that they could be a useful epidemiological early-warning indicator

The presence of CT values in each gene detected were the only values subjected to statistical analysis. Therefore, the frequency of absent CT values was not calculated. The coexistence of each gene was compared to test the significance difference on its CT values between N genes, ORF, RDRP and E genes.

The N gene (mean CT value= 28.1617) was compared the CT values of the ORF, RDRP and E genes which showed no significant difference based on their p values 0.7967 (mean CT value= 28.1687), 0.1470 (mean CT value= 29.0618), and 0.0931 (mean CT value= 27.3298), respectively.

The ORF gene was detected among 744 patients (mean CT value= 28.8603) was compared the CT values of the same frequency of 744 N and 12 RDRP genes which also showed no significant difference based on their p values *0.008* (mean CT value= 26.9801) and 0.0938* (mean CT value= 28.7000), respectively.

Coexistence of the RDRP gene-detected patients with N genes, ORF, and E gene showed variable results when their CT values were compared. The 213 RDRP gene-detected samples were significantly different with the 196 N gene-positive (0.0014) patients (p value=0.0014) and also with 188 E gene-detected (p value 0.0015) patients. However, CT values when compared to ORF, this showed that there was no significant difference between the two steps of study population as shown in Table 7.

Lastly, CT values of 188 E genes were shown to be significantly different relative to the same frequency of RDRP genes (188, p value=0.0001) while there was no significant difference when compared to 188 N gene CT values (p value= 0.0720).

This study aimed at providing a city-wide overview of the plate-based RT-PCR for SARS COV-19 during the study period. Patients covered were all admitted and confirmed with COVID-19 infection. Consolidated data were analyzed based on the available target population among different age groups, between sexes, different Cycle threshold values ranges and specific gene detection per RT-PCR. Trends and correlations were congruent to some earlier studies during the pandemic. A higher number of samples from these admitted patients represents a strength point of the study and a common laboratory where samples were analyzed. These raw data gatherers were useful for epidemiological indicators on a city-wide point of view to derive an strategic plan on a smaller scale. Further studies are highly recommended among these patients which the study was currently limited to the scope based on their clinical conditions, comorbidities and origin of swab samples.

## Data Availability

All data produced in the present work are contained in the manuscript.

## REFERENCES

Centers for Disease and Prevention. (2022). https://www.cdc.gov/coronavirus/2019-ncov/variants/variant-classifications-html. Retrieved October 13, 2022.

Cho, H., Jung, Y.H., Cho, H.B., Kim, H.T., & Kim, K.S. (2020). Positive control synthesis method for COVID-19 diagnosis by one-step real-time RT-PCR. Clin Chim Acta. Dec;511:149–153. doi: 10.1016/j.cca.2020.10.001. Retrieved October 15, 2022.

Colton, H., Ankcorn, M., Yavuz, M., Tovey, L., Cope, A., Raza, M., & Keeley, A.J., et al., (2020). Improved sensitivity using a dual target, E and RdRp assay for the diagnosis of SARS-CoV-2 infection: Experience at a large NHS Foundation Trust in the UK. J Infect. Jan;82(1):159–198. doi: 10.1016/j.jinf.2020.05.061. https://www.ncbi.nlm.nih.gov/pmc/articles/PMC7255707/. Retrieved October 14, 2022.

Kim, K. B., Choi, H., Lee, G. D., Lee, J., Lee, S., Kim, Y., Cho, S. Y., et al., (2021). Analytical and Clinical Performance of Droplet Digital PCR in the Detection and Quantification of SARS-CoV-2. Molecular diagnosis & therapy, 25(5), 617–628. https://doi.org/10.1007/s40291-021-00547-1. Retrieved October 16, 2022.

Kim, Y., Cheon, S., Jeong, H., Park, U., Ha, N. Y., Lee, J., Sohn, K. M., et al., (2021). Differential Association of Viral Dynamics With Disease Severity Depending on Patients’ Age Group in COVID-19. Frontiers in microbiology, 12, 712260. https://doi.org/10.3389/fmicb.2021.712260, Retrieved October 25, 2022.

National Library of Medicine. (2022). ORF1ab ORF1a polyprotein;ORF1ab polyprotein [Severe acute respiratory syndrome coronavirus 2]. National Center for Biotechnology Information. https://www.ncbi.nlm.nih.gov/gene/43740578. Retrieve November 12, 2022.

Singanayagam, A,, Patel, M., Charlett, A., Lopez, B.J., Saliba, V., Ellis, J., Ladhani, S. et al., (2020). Duration of infectiousness and correlation with RT-PCR cycle threshold values in cases of COVID-19, England. Euro Surveill. 25(32):2001483. doi: 10.2807/1560-7917.ES.2020.25.32.2001483. Erratum in: Euro Surveill. 2021 Feb;26(7): retrieved November 6, 2022.

Walker, S., Pritchard, E., House, T., Robotham, J.V., Birrell, P.J., Bell, I., & Bell, J.I., et al. (2021). COVID-19 Infection Survey team. Ct threshold values, a proxy for viral load in community SARS-CoV-2 cases, demonstrate wide variation across populations and over time. Elife. 12;10:e64683. doi: 10.7554/eLife.64683. PMID: 34250907; PMCID: PMC8282332. Retrieved October 22, 2022.

Waudby-West, R., Parcell, B.J., Palmer, C.N.A., Bell, S., Chalmers, J.D., & Siddiqui, M.K. (2021). The association between SARS-CoV-2 RT-PCR cycle threshold and mortality in a community cohort Ct values from RT-PCR tests are associated with risk of mortality in SARS-CoV-2 infection. European Respiratory Journal, 58 (1) 2100360; DOI: 10.1183/13993003.00360-2021, Retrieved August 13, 2022.

